# Performance-based physical functioning and incident IADL limitations in a cohort of community-dwelling older women

**DOI:** 10.1101/2025.05.15.25327231

**Authors:** Jennifer G. Lyons, Lauren Wise, Katie M. Applebaum, Kristine Ensrud, Lisa Fredman

## Abstract

**Background:** Limitations in activities of daily living have widespread implications for the well-being of older adults. However, the relation between performance-based physical function and self-reported functional impairment is inconsistent.

**Methods:** The cohort included 6,282 White women and 310 Black women aged 65 and older participating in the Study of Osteoporotic Fractures (SOF) from 1986 to 2010 who reported no limitations in any Instrumental Activities of Daily Living (IADL) at baseline. Approximately every two to six years, participants self-reported their physical limitations and trained interviewers assessed common measures of physical performance (i.e., usual gait speed, grip strength, and chair stand time). We used Cox proportional hazards models using age as the time scale to calculate hazard ratios between individual and summary measures of physical performance and incident IADL limitations.

**Results:** Over follow-up, 4,193 White women and 118 Black women developed IADL impairment (IR = 451.34 and 361.52 per 10,000 person-years, respectfully). Usual gait speed was associated with IADL limitations in both race cohorts (slowest gait vs. fastest gait HR: 3.83, 95% CI: 3.41 – 4.31; HR: 2.59, 95% CI: 1.42 – 4.73). For every one-point increase in summary performance score, rate of IADL limitations was lower for both White women and Black women (HR: 0.79, 95% CI: 0.78-0.80; HR: 0.87, 95% CI: 0.81 – 0.94).

**Conclusion:** In this longitudinal study, women with poorer performance in individual and summary measures of physical function had an increased rate of incident IADL limitations over follow-up compared to women with the best performance. These findings confirm previous research using cross-sectional data.

## Introduction

Functional impairment, defined as the difficulty performing activities of daily living (ADLs) and/or instrumental activities of daily living (IADLs), affects the psychological, social, and physical health of older adults ^1^. Self-reported limitations in any of the IADLs incorporate aspects of an older adult’s ability to function in everyday life, and encompasses perceived physical competence, chronic condition burden, and even depressive symptoms.^2–4^ However, the relation between performance-based physical function and self-reported functional impairment is inconsistent. Several studies have found moderate to strong associations between performance-based physical function and subsequent impairment,^1,3,5–8^ but results vary by the study cohort characteristics and the performance measures used. These inconsistent associations may result from inadequate control for age as a confounder of this association. Age is the strongest predictor of function and impairment: physical function declines with age^9–11^ and the rate of this decline is faster at older ages.^12^ As such, physical function that is not updated over time is vulnerable to misclassification over long follow-up periods, particularly among older adults. Further, impairment in activities of daily living are higher in women than in men,^4,13^ making older women an important target population to investigate the association between reported function and physical impairment. To date, previous studies have evaluated performance-based functioning as a baseline exposure^1,8,14^, and others had short follow-up time (e.g., two or three years)^7,15^ or excluded women.^15,16^ Further, no study of this association has included time-varying exposures and confounders, or age-based risk-sets to finely control for age. The current study utilizes time-dependent analysis and age-based risk sets to minimize confounding by age and misclassification of physical function over time.

In this study, we examined the association of three individual measures of lower- and upper-extremity physical functioning independently and as a summary score with the first occurrence of an IADL limitation. To account for longitudinal changes in function, interviewers reassessed the performance measures at every clinical evaluation. We hypothesized that older community-dwelling women with poorer physical function of each individual summary measure (e.g., slower gait speed, lower grip strength, slower chair stand time) over time would have a greater incidence of IADL limitations compared to those with the best physical function, and these associations would be strongest for gait speed. We also hypothesized that older women with poorer performance overall (e.g., low summary performance score) over time would have a greater incidence of IADL limitations compared to those with the best physical function. Furthermore, these time-varying associations would be stronger than those using only baseline measures of physical function.

## Methods

### Data Collection and Analytic Sample

The SOF sample includes 9,704 women aged 65 or older who were recruited between 1986 and 1988 from population-based listings in four areas of the United States: Baltimore County, MD; Minneapolis, MN; Portland, OR; and the Monongahela Valley, PA. Women were excluded if they could not walk without assistance or had a history of bilateral hip replacement. Although Black women were initially excluded because of their low incidence of hip fracture, 662 Black women who met the same inclusion criteria were enrolled during 1996-97. Approximately every two to six years, SOF participants had a comprehensive clinical evaluation to assess physical and cognitive health. All participants provided written informed consent, and the Institutional Review Boards of each study site approved the study protocol.

At Visit 1 (1986-1987), 6,296 White women (65% of the sample) reported no limitations in any Instrumental Activities of Daily Living (IADLs). At their baseline clinical evaluation (Visit 6, 1997-1998), 320 Black women (48% of the sample) reported no limitations in any IADLs. Participants contributed person-time from the date of their first clinical visit to either the date of incident IADL limitations, withdrawal from SOF, death, or end of the follow-up period on December 31, 2010, whichever came first. White women contributed a maximum of nine visits and Black women contributed a maximum of four visits through December 31, 2010, for a maximum of 24 years and 14 years of follow-up, respectively. The analytic sample consisted of 6,282 White women and 310 Black women who reported no IADL difficulties and completed all three measures of physical performance at baseline.

### Outcome: IADL Impairment

Interviewers asked participants to describe their ability to complete five instrumental activities of daily living tasks^17,18^ at every interview. Participants reported whether they could complete each of the tasks with or without difficulty: walking 2-3 blocks outside on level ground, climbing 10 steps without stopping/resting, preparing own meals, doing heavy housework, and shopping. The resulting dichotomous response variables (yes versus no) were then summed to create a summary IADL score (range: 0-5), with higher values indicating greater limitations. We defined incident IADL limitations at the first SOF follow-up interview at which a woman reported difficulty in any one of the five IADL tasks.

### Predictors: Individual Performance Measures and Summary Performance Score

Trained interviewers assessed three common measures of physical performance: usual gait speed, grip strength, and chair stand time. Participants were asked to walk their usual gait speed (meters/second, m/s) over a straight six-meter course and speed was averaged over two timed trials.^19^ Faster speeds indicated better performance (i.e., higher values for m/s). In prior studies, gait speed was found to have good test-retest reliability.^19,20^ Maximum isometric grip strength (kilograms, kg) was measured in both hands with a dynamometer (Preston Grip dynamometer, Takei Kikikogyo, Tokyo, Japan). Grip strength was tested twice in each hand and averaged over the four trials. If participants could not complete the test in one hand, the two trials from the other hand were averaged. Stronger grip strength indicated better performance (i.e., higher values for kilograms, kg). Earlier studies in SOF found that weaker grip strength was associated with impaired function.^19^ Chair stand time (seconds) was measured as the time it took participants to stand up from a seated position in a straight-back chair five times, with their arms folded across their chest.^21,22^ Faster chair stand time indicated better performance (i.e., fewer seconds to complete). The chair stand test has good test-retest reliability in older, community-dwelling adults.^23^

We calculated a summary performance score based on the three component measures by calculating race-specific sample-based quartiles for each individual summary performance score. Participants who attempted but were unable to complete a measure were excluded from the quartiles for that measure and scored as 0 (range: 0 – 4).^1^ Quartiles for each component measure were summed to create a composite summary score (0-12). Higher scores indicated better performance. The summary performance score was evaluated as both a categorical variable,^1^ and a continuous variable.

### Sociodemographic and Health Characteristics

Sociodemographic variables included the respondent’s age, marital status (married versus other), highest level of education (> 12 years versus ≤ 12 years), and SOF study site. Health status variables included body mass index (BMI, weight in kilograms/height in meters^2^, categorized according to standard cut points as <18.5, 18.5-24.9, 25-30, ≥30 km/m^2^) based on measured weight and height; and smoking status (never, past, current).

We calculated a multimorbidity score by summing five domains of medical conditions.^24^ Participants reported whether they had ever been diagnosed, or had been diagnosed since their last examination, with any of 11 medical conditions (i.e., osteoarthritis, osteoporosis, rheumatoid arthritis, cancer, stroke, myocardial infarction, congestive heart failure, other heart disease, hypertension, type 2 diabetes, and chronic obstructive pulmonary disease) that were further classified into five health domains (i.e., cardiovascular and respiratory, metabolic, cancer, immune, and musculoskeletal). The five health domains were summed and operationalized as a three-level categorical variable (no conditions in any domain, conditions in a single domain, and conditions in two or more domains).

### Statistical Analyses

To accommodate race-dependent differences in the enrollment period, we stratified the regression analyses by race. We evaluated the relationship between physical function and IADL limitations in two sets of models, one using baseline measurements only and one using time-dependent performance measures and covariates.

In descriptive analyses we compared distributions of baseline sociodemographic and health characteristics in the total analytic sample and stratified by race. We compared means and standard deviations for continuous variables, calculated frequencies and proportions for dichotomous and categorical variables, and estimated crude incidence rates of the association between each physical performance measure and first IADL limitation.

In inferential analyses, we used Cox proportional hazards models using age as the time scale to calculate age-adjusted and adjusted hazard ratios (HR and aHR) and 95% confidence intervals (CI) between physical performance (separately for the individual measures and summary score) and incident IADL limitations. We used the Andersen-Gill data structure^25^ to accommodate time-dependent covariates and delayed entry, a consequence of using age as the time scale. Covariates were included in the model if they were established risk factors for poor physical performance or IADL limitations, if they were associated with any physical performance measure or IADL limitations in the current sample, or if their inclusion in the proportional hazards model meaningfully changed the HR of the association between poor physical performance and IADL limitations by 10% or more. The final adjusted models included education, marital status, smoking status, BMI, multimorbidity score categories, and SOF clinical center. We assessed departures from proportional hazards by comparing models with and without interaction terms between physical performance quartiles and age (median cut point of <75 versus ≥75 years of age) using the likelihood ratio test, as well as by examining the log-negative-log survival curves of these models.

All analyses were conducted using SAS software (Statistical Analysis System, version 9.4, Cary, North Carolina). All statistical tests were two-sided and used 95% confidence intervals.

## Results

### Sample Characteristics

At the baseline clinical examination, the mean age of the 6,282 White women was 71.05 years (SD=4.91), 51% were married, and 80% had graduated high school. More than a third of the cohort reported medical conditions in at least one domain (37.71%, Table 1). Two-thirds of the sample (66.59%) experienced incident IADL limitations over the follow-up period. Over the entire follow-up period, 3,593 (57.20%) women died and 678 (10.79%) terminated from SOF (data not shown).

**Table 1.** Baseline demographic, health, and performance characteristics of 6,592 SOF participants with no IADL IADL limitations at baseline.

|  | Total sample<br>N=6,592 |  | White women<br>N=6,282 |  | Black women<br>N=310 |  |
| --- | --- | --- | --- | --- | --- | --- |
| Age, mean (sd) | 71.22 | (4.95) | 71.05 | (4.91) | 74.76 | (4.45) |
| High school education, n (%) | 5,274 | (80.01) | 5,040 | (80.23) | 234 | (75.48) |
| Married, n (%) | 3,294 | (49.97) | 3,207 | (51.05) | 87 | (28.06) |
| Smoking status, n (%) |  |  |  |  |  |  |
| Never smoker | 4,062 | (61.62) | 3,868 | (61.57) | 194 | (62.58) |
| Ever smoker | 1,891 | (28.69) | 1,796 | (28.59) | 95 | (30.65) |
| Current smoker | 615 | (9.33) | 594 | (9.46) | 21 | (6.77) |
| Short Mini Mental Status Exam, mean (sd) | 24.71 | (1.66) | 24.78 | (1.60) | 23.37 | (2.27) |
| BMI, mean (sd) | 26.01 | (4.18) | 25.85 | (4.09) | 29.34 | (4.71) |
| <18.5, n (%) | 109 | (1.65) | 86 | (1.37) | 23 | (7.42) |
| 18.5-24.9, n (%) | 2,927 | (44.40) | 2,872 | (45.72) | 55 | (17.74) |
| 25-29.9, n (%) | 2,495 | (37.85) | 2,374 | (37.79) | 121 | (39.03) |
| ≥30, n (%) | 1,061 | (16.10) | 950 | (15.12) | 111 | (35.81) |
| Osteoarthritis, n (%) | 1,790 | (27.15) | 1,769 | (28.16) | 21 | (6.77) |
| Myocardial infarction, n (%) | 8 | (0.12) | 0 | (0.00) | 8 | (2.58) |
| Hypertension, n (%) | 1,219 | (18.49) | 1,109 | (17.65) | 110 | (35.48) |
| Diabetes, n (%) | 214 | (3.24) | 185 | (2.94) | 155 | (50.00) |
| COPD, n (%) | 16 | (0.24) | 0 | (0.00) | 16 | (5.15) |
| Multimorbidity score, n (%) |  |  |  |  |  |  |
| No domains | 4,076 | (61.83) | 3,913 | (62.29) | 163 | (52.58) |
| 1 domain | 1,586 | (24.06) | 1,490 | (23.72) | 96 | (30.97) |
| ≥ 2 domains | 930 | (14.11) | 879 | (13.99) | 51 | (16.45) |
| Gait speed, (m/second) mean (sd) | 1.06 | (0.20) | 1.07 | (0.20) | 0.92 | (0.17) |
| Grip strength (kg), mean (sd) | 21.50 | (4.13) | 21.55 | (4.08) | 20.32 | (4.89) |
| Chair stand time (s), mean (sd) | 11.39 | (3.37) | 11.29 | (3.30) | 13.53 | (4.08) |
| Summary performance score, mean (sd) | 8.43 | (2.20) | 8.46 | (2.20) | 7.92 | (2.25) |

Note. BMI – Body Mass Index; COPD – Chronic Obstructive Pulmonary Disease

Black women were older and had more medical conditions than White women. At the baseline clinical examination, the mean age of the 310 Black women was 74.76 years (SD=4.45), 28% were married, and 75% had graduated high school. Almost half of the cohort reported medical conditions in at least one domain (47.42%, Table 1). About a third of the sample experienced incident IADL limitations over follow-up (36.77%), which likely was a much lower proportion than the White cohort due to a shorter follow-up time. Over the entire follow-up period, 60 (19.35%) women died and 34 (10.9%) terminated from SOF (data not shown).

### Physical Performance

Distributions of gait speed and chair stand times were different between the two cohorts, with Black women performing more poorly than White women (Table 1-A1). For example, only the fastest Black women (quartile four) walked faster than the clinically relevant gait speed cut point of 1.0 m/s.^20^ However, in both cohorts, the lowest grip strength quartile cut point was the same (16.50 kg) and closely corresponded to cut points that predict IADL limitations in women over 65 years of age.^26^

At baseline, mean gait speed was 1.07 m/s (SD=0.20) and 35.28% of White women walked slower than 1.0 m/s. Mean grip strength was 21.55 kgs (SD=4.89) and mean chair stand time was 11.29 seconds (SD=3.30, Table 1). At the baseline visit, the largest proportion of White women were in the highest quartile of gait speed (37.23%) and grip strength (42.93%), and the second highest quartile of chair stand time (26.23%) compared to the lower quartiles (Table 2). Mean summary performance score was 7.20 (SD=1.48, Table 1).

**Table 2.** Baseline performance measure characteristics among 6,592 SOF participants with no IADL disabilities at baseline.

|  | Total |  | White women |  | Black women |  |
| --- | --- | --- | --- | --- | --- | --- |
|  | n | (%) | n | (%) | n | (%) |
| Performance Measure Quartiles |  |  |  |  |  |  |
| Gait Speed |  |  |  |  |  |  |
| Q1 | 903 | (13.70) | 861 | (13.71) | 42 | (13.55) |
| Q2 | 1,350 | (20.48) | 1,268 | (20.18) | 82 | (26.45) |
| Q3 | 1,903 | (28.87) | 1,814 | (28.88) | 89 | (28.71) |
| Q4 | 2,436 | (36.95) | 2,339 | (37.23) | 97 | (31.29) |
| Grip strength |  |  |  |  |  |  |
| Q1 | 672 | (10.19) | 606 | (9.65) | 66 | (21.29) |
| Q2 | 1,166 | (17.69) | 1,092 | (17.38) | 74 | (23.87) |
| Q3 | 1,964 | (29.79) | 1,887 | (30.04) | 77 | (24.84) |
| Q4 | 2,790 | (42.32) | 2,697 | (42.93) | 93 | (30.00) |
| Chair stand time |  |  |  |  |  |  |
| Q1 | 1,564 | (23.73) | 1,490 | (23.72) | 74 | (23.87) |
| Q2 | 1,722 | (26.12) | 1,639 | (26.09) | 83 | (26.77) |
| Q3 | 1,721 | (26.11) | 1,648 | (26.23) | 73 | (23.55) |
| Q4 | 1,585 | (24.04) | 1,505 | (23.96) | 80 | (25.81) |
| Summary Score |  |  |  |  |  |  |
| 3 | 104 | (1.58) | 99 | (1.58) | 5 | (1.61) |
| 4 | 225 | (3.41) | 205 | (3.26) | 20 | (6.45) |
| 5 | 399 | (6.05) | 376 | (5.99) | 23 | (7.42) |
| 6 | 600 | (9.10) | 561 | (8.93) | 39 | (12.58) |
| 7 | 821 | (12.45) | 774 | (12.32) | 47 | (15.16) |
| 8 | 1,015 | (15.40) | 969 | (15.43) | 46 | (14.84) |
| 9 | 1,098 | (16.66) | 1,055 | (16.79) | 43 | (13.87) |
| 10 | 1,039 | (15.76) | 995 | (15.84) | 44 | (14.19) |
| 11 | 815 | (12.36) | 788 | (12.54) | 27 | (8.71) |
| 12 | 476 | (7.22) | 460 | (7.32) | 16 | (5.16) |
| Continuous, mean (sd) | 8.18 | (2.26) | 8.20 | (2.26) | 7.71 | (2.26) |

Black women had poorer performance across all measures at baseline than White women. At baseline, mean gait speed was 0.92 m/s (SD=0.17) and 68.71% of Black women walked slower than 1.0 m/s. At the baseline visit, mean grip strength was 20.32 (SD=4.89) and mean chair stand time was 13.53 seconds (SD=4.08, Table 1). The largest proportion of Black women were in the highest quartile of gait speed (31.29%), grip strength (30.00%), and the second lowest quartiles of chair stand time (26.77%), noting that quartile cut points for all physical performance measures were lower in this cohort than for the White cohort (Table 2). Mean summary performance score was 7.35 (SD=1.58, Table 1).

### Physical Performance and Incident IADL Impairment

Over 92,901 years of person-time, 4,193 White women developed an IADL limitations (IR = 451.34 per 10,000 person-years). In both age-adjusted and fully adjusted analyses of baseline gait speed, there was a monotonic increase in rate of incident IADL limitations across gait speed quartiles (Q1 vs. Q4 adjusted HR: 1.65, 95% CI: 1.48 – 1.84, Table 3). This association was stronger in time-dependent analyses (Q1 vs. Q4 aHR: 3.83, 95% CI: 3.41 – 4.31, Table 4). A similar pattern was found with grip strength and chair stand time. Women in the lowest quartile of each measure had a greater rate of incident IADL limitations than those in the highest quartile, and these associations were stronger in time-dependent analyses (grip strength Q1 vs. Q4 aHR: 2.15, 95% CI 1.91 – 2.41; chair stand Q1 vs. Q4 aHR: 2.69, 95% CI: 2.42 – 2.99, Table 4) compared to baseline analyses (grip strength Q1 vs. Q4 aHR: 1.49, 95% CI 1.33 -1.68; chair stand Q1 vs. Q4 aHR: 1.65, 95% CI: 1.50 – 1.80, Table 4). While patterns of increased rate of IADL limitations across performance quartiles remained similar between the baseline and time-dependent analyses, the estimated rate of incident IADL limitations was attenuated in the baseline analyses.

**Table 3.**
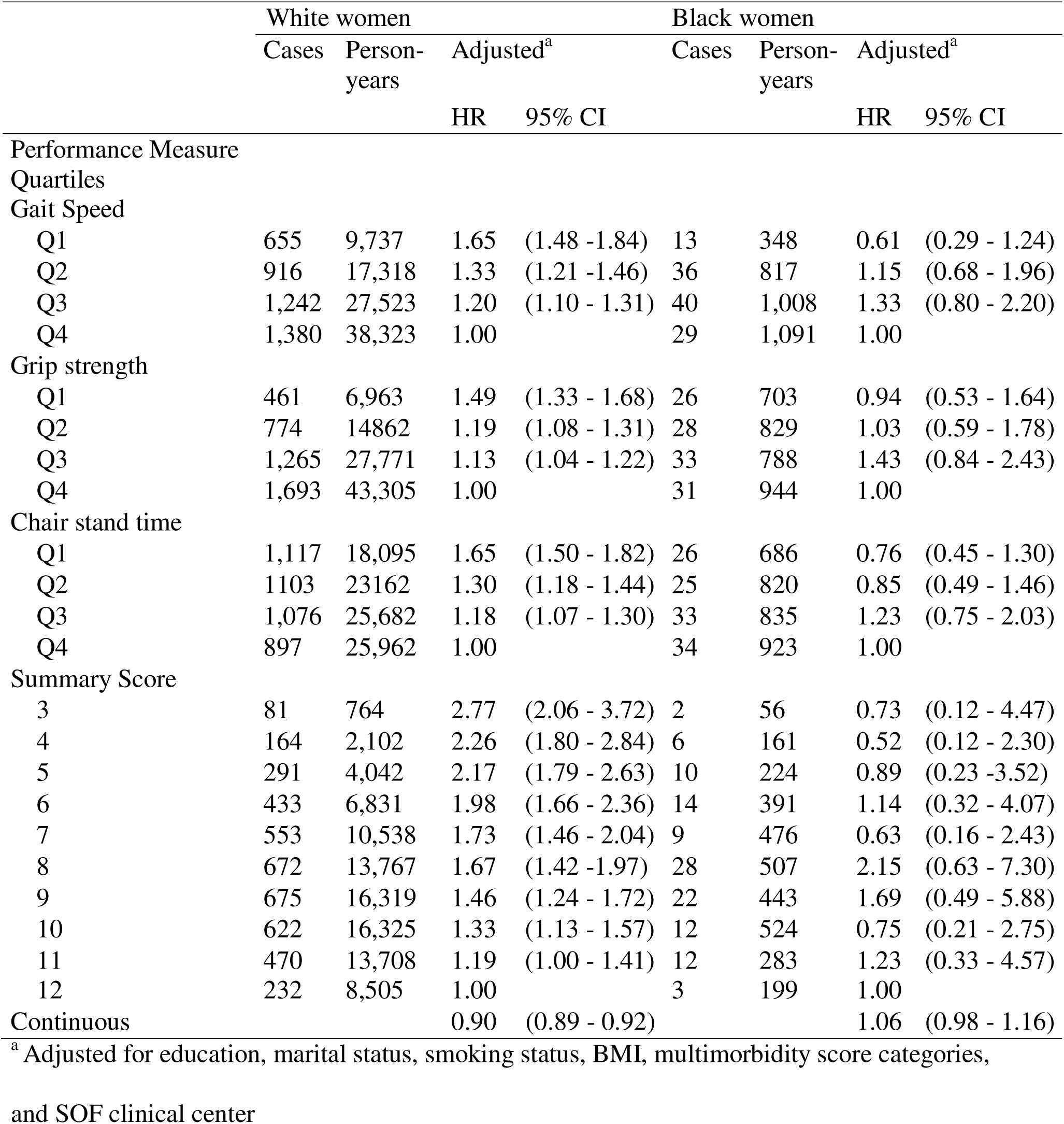
Associations between baseline physical performance quartiles and summary score with incident IADL IADL limitations, by race, among 6,592 SOF participants.

**Table 4.** Associations between time-dependent physical performance quartiles and summary score with incident IADL IADL limitations, by race, among 6,592 SOF participants.

|  | White women |  |  |  | Black women |  |  |  |
| --- | --- | --- | --- | --- | --- | --- | --- | --- |
|  | Cases | Person-years | Adjusted<sup>a</sup> |  | Case | Person-years | Adjusted<sup>a</sup> |  |
|  |  |  | HR | 95% CI |  |  | HR | 95% CI |
| Gait Speed, m/s |  |  |  |  |  |  |  |  |
| Unable | 14 | 64 | 6.55 | (3.81 - 11.24) | 2 | 8 | 3.72 | (0.79 - 17.49) |
| Q1 | 1,759 | 19,890 | 3.83 | (3.41 - 4.31) | 50 | 546 | 2.59 | (1.42 - 4.73) |
| Q2 | 1,021 | 22,323 | 2.03 | (1.79 - 2.30) | 33 | 833 | 1.36 | (0.73 - 2.54) |
| Q3 | 836 | 25,236 | 1.53 | (1.35 - 1.74) | 17 | 886 | 1.25 | (0.62 - 2.50) |
| Q4 | 563 | 25,388 | 1.00 |  | 16 | 991 | 1.00 |  |
| Grip strength, kg |  |  |  |  |  |  |  |  |
| Unable | 49 | 586 | 2.19 | (1.61 - 2.99) | 6 | 66 | 1.81 | (0.70 - 4.67) |
| Q1 | 1,563 | 21,613 | 2.15 | (1.91 - 2.41) | 38 | 697 | 1.50 | (0.86 - 2.62) |
| Q2 | 1,050 | 22,194 | 1.54 | (1.37 - 1.73) | 27 | 905 | 1.01 | (0.55 - 1.84) |
| Q3 | 894 | 24,586 | 1.23 | (1.09 - 1.39) | 25 | 772 | 1.18 | (0.65 - 2.16) |
| Q4 | 637 | 23,922 | 1.00 |  | 22 | 824 | 1.00 |  |
| Chair stand time, s |  |  |  |  |  |  |  |  |
| Unable | 13 | 125 | 2.14 | (1.20 - 3.81) | 0 | 10 | 0.00 | (0.00 - 0.00) |
| Q1 | 1,597 | 20,453 | 2.69 | (2.42 - 2.99) | 32 | 728 | 1.28 | (0.72 - 2.30) |
| Q2 | 1,118 | 23,292 | 1.82 | (1.63 - 2.03) | 33 | 821 | 1.27 | (0.72 - 2.26) |
| Q3 | 867 | 24,289 | 1.35 | (1.20 - 1.52) | 32 | 857 | 1.42 | (0.80 - 2.52) |
| Q4 | 628 | 24,742 | 1.00 |  | 21 | 848 | 1.00 |  |
| Summary Score |  |  |  |  |  |  |  |  |
| 0 | 17 | 168 | 5.66 | (3.15 - 10.16) | 1 | 19 | 0.64 | (0.06 - 6.65) |
| 1 | 52 | 263 | 9.21 | (5.83 - 14.53) | 2 | 6 | 2.84 | (0.43 - 18.76) |
| 2 | 162 | 826 | 11.15 | (7.77 - 16.00) | 5 | 50 | 0.78 | (0.16 - 3.87) |
| 3 | 515 | 4,056 | 8.26 | (5.95 - 11.47) | 21 | 137 | 2.34 | (0.66 - 8.26) |
| 4 | 576 | 6,193 | 6.11 | (4.41 - 8.46) | 12 | 217 | 0.79 | (0.21 - 3.01) |
| 5 | 599 | 8,608 | 4.74 | (3.43 - 6.56) | 15 | 257 | 1.03 | (0.27 - 3.87) |
| 6 | 533 | 10,448 | 3.53 | (2.55 - 4.88) | 15 | 335 | 1.68 | (0.47 - 5.98) |
| 7 | 533 | 12,600 | 2.85 | (2.06 - 3.95) | 12 | 448 | 0.63 | (0.17 - 2.33) |
| 8 | 424 | 12,843 | 2.25 | (1.62 - 3.13) | 14 | 548 | 0.55 | (0.15 - 2.01) |
| 9 | 339 | 13,390 | 1.68 | (1.20 - 2.35) | 11 | 455 | 0.66 | (0.18 - 2.43) |
| 10 | 261 | 11,228 | 1.49 | (1.06 - 2.10) | 3 | 385 | 0.28 | (0.06 - 1.45) |
| 11 | 120 | 8,011 | 1.05 | (0.72 - 1.53) | 4 | 241 | 0.63 | (0.14 - 2.91) |
| 12 | 62 | 4,267 | 1.00 |  | 3 | 166 | 1.00 |  |
| Continuous |  |  | 0.79 | (0.78 - 0.80) |  |  | 0.87 | (0.81 - 0.94) |

<sup>a</sup> adjusted for education, marital status, smoking status, BMI, multimorbidity score categories,
and SOF clinical center

Across highest to lowest summary performance scores there was a clear pattern of greater rate of incident IADL limitations, although as with the individual performance measures these rates were lower in the baseline analysis compared to the time-dependent analysis (baseline score 3 vs. score 12 aHR: 2.77, 95% CI: 2.06 – 3.72; time-dependent score 3 vs. score 12 aHR: 8.26, 95% CI 5.95 – 11.47, Tables 3 and 4). For every one-point increase in summary performance score, rate of IADL limitations was 10% lower in baseline analyses (HR: 0.90, 95% CI: 0.89 – 0.92) and 21% lower in time-dependent analyses (HR: 0.79, 95% CI: 0.78-0.80).

Over 3,264 years of person-time, 118 Black women developed an IADL limitations (IR = 361.52 per 10,000 person-years). Similar to the White cohort, there was a monotonic relation between women with slower gait speed quartile and increased rate of IADL limitations, although the small number of cases and short follow-up time led to imprecise estimates that often included the null value. These associations were less precise using baseline measurements only (baseline Q1 vs. Q4 aHR: 0.61, 95% CI: 0.29 – 1.24; time-dependent Q1 vs. Q4 aHR: 2.59, 95% CI: 1.42 – 4.73, Tables 3 and 4).

Results were less clear across grip strength quartiles for both the baseline and time-dependent analyses (baseline Q1 vs. Q4 aHR: 0.94, 95% CI: 0.53 – 1.64; time-dependent Q1 vs. Q4 aHR: 1.50, 95% CI: 0.86 – 2.62, Tables 3 and 4). Results across chair stand time quartiles were inconclusive: there were no events in the lowest quartile and slight, but imprecise, increased rates of IADL IADL limitations in women with the slowest chair stand time in the time-dependent analysis but not the baseline analysis (baseline Q1 vs. Q4 aHR: 0.76, 95% CI: 0.45 – 1.30; time-dependent Q1 vs. Q4 aHR: 1.28, 95% CI 0.72 – 2.30, Tables 3 and 4). Results among all performance measure quartiles were imprecise in the Black cohort due to small number of events and short follow-up time. This pattern was similar to that observed in the White cohort, even though follow-up time was shorter for Black women.

Across the 12 summary performance categories, there was no clear pattern of greater rate of IADL limitations with lower summary scores in either the baseline analyses or the time-dependent analysis. This is likely due to small numbers in all categories: there was 1 participant in the highest category (score = 12) and 2 in the second lowest category (score = 1), which is the lowest category that included a successfully completed performance measure. However, continuous summary performance score showed a similar pattern of decreased rate of IADL limitations with increasing score as was seen in the White women, but only for time-dependent analyses. In baseline analyses, Black women had a 6% increased rate of IADL limitations for every one-point increase in score, but this association was imprecise (HR: 1.06, 95% CI: 0.98 – 1.16). In time-dependent analyses, rate of IADL limitations was 13% less for every one-point increase in score (HR: 0.87, 95% CI: 0.81 – 0.94).

## Discussion

In this longitudinal study of community-dwelling older women, women with poorer performance in individual measures of gait speed, grip strength, and chair stand speed, as well as poorer performance in a summary measure, had an increased rate of incident IADL limitations over the follow-up period compared to women with the best performance. These associations remained after adjusting for confounders and were stronger among White women compared with Black women. These findings support the initial hypotheses that women with poorer performance would have an increased rate of IADL limitations, and confirm previous research using cross-sectional data^1,14^ and shorter follow-up periods.^15^ However, while results in the Black cohort suggest a similar relation, the small number of events limited the conclusions that could be made from these associations.

The observed relations between the individual measures of physical performance and the summary score were stronger in the time-dependent analyses than those limited to baseline measurements only. Previous research has suggested that the predictive ability of these types of summary performance scores deteriorates after six years.^27^ This is likely due to exposure misclassification that biased the baseline-only results toward the null. Physical function is known to decrease with age and is expected to change over time in a longitudinal epidemiologic study with such a long follow-up period. As such, it is unlikely that baseline analyses accurately captured a participant’s true exposure status when they experienced incident IADL limitations. Thus, time-dependent analyses more precisely measure a participant’s true functional status and are more appropriate to use for analyses with long follow-up.

The summary performance score evaluated in the current study included measures of both lower and upper body strength and function, and therefore was a measure of whole body physical function. Previous studies of physical function have concluded that the association between gait speed alone and IADL limitations was similar to that of summary performance scores of lower extremity function,^27,28^ while grip strength was nearly as strongly associated with IADL limitations as a summary performance score of upper extremity function.^29^ However, these studies utilized baseline measurements of physical performance only, and none evaluated lower- and upper-extremity function together in a single summary scale. In the current study we found that the summary performance score, when measured longitudinally, was a stronger predictor of incident IADL limitations than any of the individual components and was also able to distinguish gradations of physical function better than any individual component. Given these findings, there is added benefit of including several measures of physical performance in a summary score, particularly when taking measurements longitudinally.

In both race cohorts, gait speed was the strongest predictor of incident IADL limitations. This finding confirms previous research that suggests gait speed is a strong predictor of IADL limitations and other adverse health outcomes in older adults.^1,20,30–32^ Gait speed incorporates a variety of factors relating to physical function, including motor control,^33^ muscle strength,^34,35^ and musculoskeletal condition;^36^ it also is associated with health characteristics that are not related to physical capabilities, such as cognitive function^37,38^ and comorbidities.^39^ Gait speed may be a mediator on the causal pathway from poor health to IADL limitations, and can be considered a proxy measure of general health^40,41^ that affects risk of future IADL limitations.

Updating gait speed measurements over time allow to better capture current physical function, which resulted in even strong associations between gait speed and IADL limitations.

There are several potential limitations to the current study. Notably, there was a concern about outcome misclassification, as IADL limitations is not a simple or straightforward diagnosis. Over a lengthy follow-up period, as is the case in SOF, participants may report IADL limitations at one interview but not report that same limitation at subsequent interviews. This improvement may be real (i.e., recovery after a hip replacement or illness) or an incorrect perception of their abilities after acclimating to their IADL limitations. However, research suggests that a very small proportion of those who report increased IADL limitations improve,^42^ and fewer than 20% of SOF participants who reported a limitation over follow-up never reported a limitation again. The sensitivity analysis which included women with prevalent IADL limitations addressed this issue; results from these sensitivity analyses demonstrated that potential outcome misclassification was minimal. Further, any misclassification that did occur was unlikely to be dependent given that the exposure was based on self-report and the performance-based measures were administered at the clinic by a trained interviewer.

White women contributed more person-years than Black women, which allowed for the observation of fewer events in this group. As such, rates were imprecise for the Black cohort, though there was still suggestion of the general pattern that women in lower quartiles of all three physical performance measures had higher rates of IADL limitations than those in the highest quartile. We expected that the larger sample size in the sensitivity analyses would allow for more events to be observed in the Black cohort, which would improve the precision of the results. However, this was not the case. It is not clear whether the lack of association between the individual performance measures or the summary performance score was due to small numbers of events, insufficient follow-up time, or a true relationship. However, results for gait speed were an exception to this observation. These results suggested that there was an association between slower gait speed and greater rates of IADL limitations in the Black cohort.

Another concern was possible dependent misclassification of gait speed, which may have strong effects on the association between both the individual measure of function as well as the summary measure, which included gait speed quartiles. While participants who completed the SOF questionnaires at the clinic all complete a six-meter walking course, participants who opted to complete an interview at their home (often because poor health made it difficult for them to travel to the clinic) may not have had that length of unobstructed walking space available, and may have had data only on shorter course lengths (i.e., 2-meter or 3-meter). Shorter course lengths likely over-estimate the amount of IADL limitations in a population^43^ and in this sample, those who completed the shorter course lengths were more likely to be in poor health, and were more likely to develop a IADL limitations, than those who completed the longer course length. This relationship between poorer health and shorter course length may bias the association between gait speed and IADL limitations toward the null. However, less than 3% of all the gait speed measurements over follow-up were completed on course less than 6 meters long, and all baseline gait speeds were measured on a 6-meter course, so this bias should be minimal.

In addition, the poorest performers, those who attempted but were unable to complete at least one physical performance measure, contributed the least amount of follow-up time. It is possible that these women were poor performers because of their overall health, which would not only increase their likelihood of developing IADL limitations but would also be related to their censoring (discontinuation from SOF or death). If censoring prevented the observation of incident IADL limitations, this might have resulted in underestimating the association between performance and IADL limitations. In several instances, for example the regression results for the summary performance score in both race cohorts, the lowest performers (score = 0) had a lower adjusted risk of incident IADL limitations compared to the highest performers (score = 12). These results may be attenuated, though further investigation via quantitative bias analysis is needed to determine the extent.

Nonetheless, this study had many strengths. The sample was relatively large with a long follow-up period (18 years for White women and 13 years for Black women) allowing enough time to accrue to capture many events, particularly in the White cohort. The exposure utilized sample-based cut points to categorize physical performance, which replicated previous studies^1^ and accommodated the distributions in function in this cohort that are not accurately captured with clinical cut points^20,44,45^ used for other populations. Unlike previous studies of the association between physical performance and IADL limitations, which used only baseline measurements and cross-sectional analyses^1,14^, the current study used time-varying measures of physical performance to predict incident IADL limitations longitudinally. In addition, physical performance deficits increase with age,^9,10,12^ and allowing these performance measures to vary over time reduced the likelihood of exposure misclassification. Accounting for variability over time also captured improvements in function which, while not common in this population of older women, would bias the results toward the null if not accounted for.

Similarly, as physical performance and IADL limitations are so closely linked to age, using age as the time scale instead of time in study allows for more precise control for age-related confounding. In the current study using the Andersen-Gill data structure, participants were included in the analysis in age-based risk sets; if an event occurred at an age at which a participant was under observation, that participant would contribute person-time for that age. As such, risk of IADL limitations was only compared among women of the same age. This reduced, and may have even eliminated, confounding by age-related factors that were not collected or controlled for in the analysis. Further study is needed to evaluate potential time-dependent confounding and potential effect measure modification by age that is not addressed with the current analysis.

In summary, in this longitudinal study of community-dwelling older women, physical performance measured at baseline and over time was associated with incident IADL limitations. Utilizing time-dependent physical performance measures yielded higher, and less misclassified, risks across performance categories. Objective measures of physical performance, particularly gait speed, are associated with a variety of health and functional outcomes. Taken together, these results suggest that poorer physical performance is likely on the causal pathway of poor health to IADL limitations.

## Data Availability

These data are from the Study of Osteoporotic Fractures, a longitudinal epidemiologic study of older women with 20 years of follow-up. These data are not publicly available.

## Acknowledgments

The authors would like to thank Timothy Heeren, PhD for his contribution to this work and manuscript.

---

**Figure 1:**
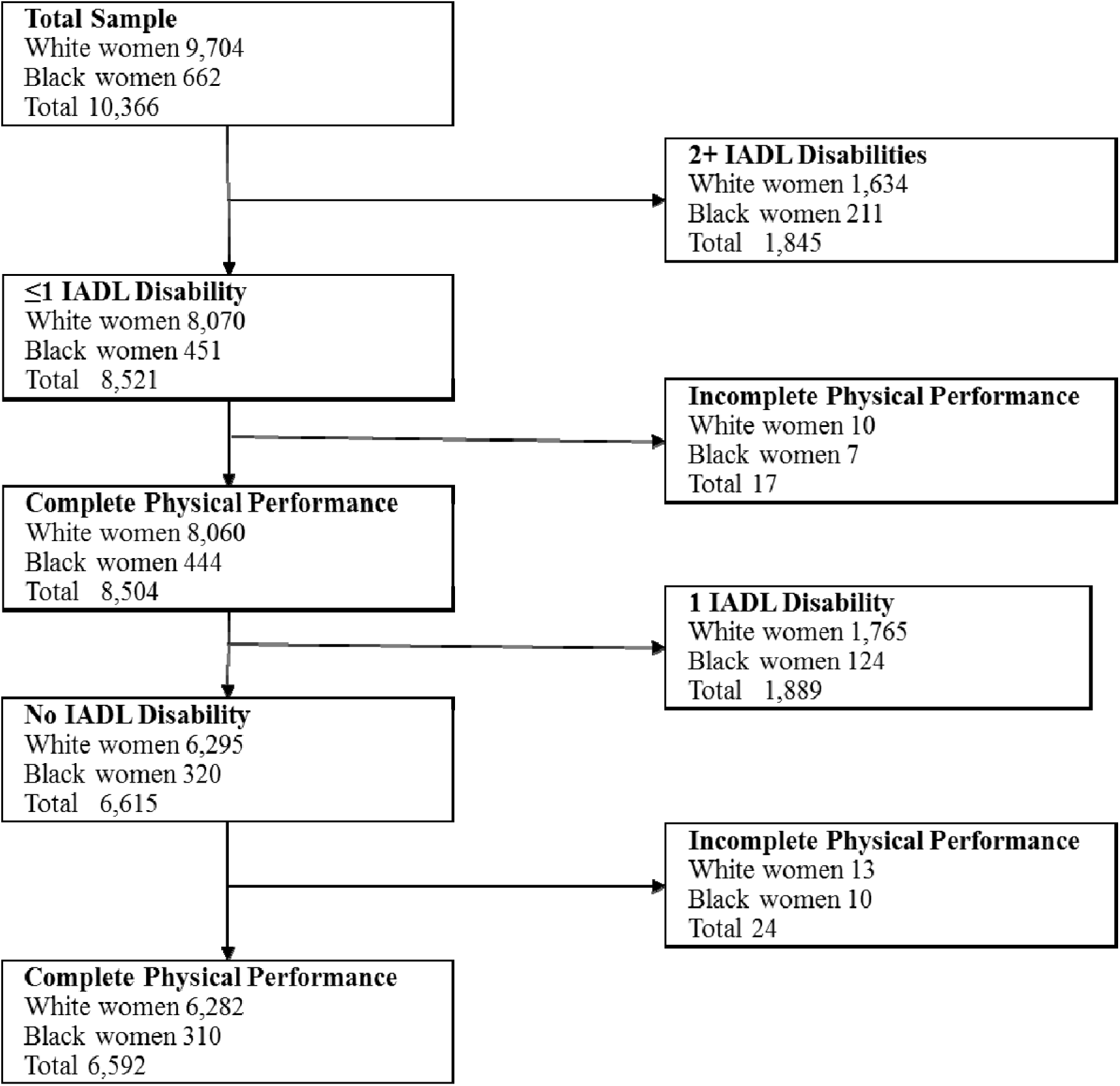
Selection of analytic sample

**Table 1-A1:** Quartile cut points for each individual performance measure, by race cohort.

|  | White Women | Black Women |
| --- | --- | --- |
| Gait Speed (m/s) |  |  |
| Quartile 1 | <0.86 | <0.75 |
| Quartile 2 | 0.86 - 0.98 | 0.75 - 0.87 |
| Quartile 3 | 0.99 - 1.12 | 0.88 - 0.99 |
| Quartile 4 | ≥1.13 | ≥1.00 |
| Grip Strength (kg) |  |  |
| Quartile 1 | <16.50 | <16.50 |
| Quartile 2 | 16.50 - 19.49 | 16.50 - 19.99 |
| Quartile 3 | 19.50 - 22.49 | 20.00 - 22.99 |
| Quartile 4 | ≥22.50 | ≥23.00 |
| Chair Stand time (sec) |  |  |
| Quartile 1 | ≥13.10 | ≥16.00 |
| Quartile 2 | 10.80 - 13.09 | 13.10 - 15.99 |
| Quartile 3 | 9.10 - 10.79 | 10.70 - 13.09 |
| Quartile 4 | <9.10 | <10.70 |

## References

1. Guralnik JM, Simonsick EM, Ferrucci L, et al. A short physical performance battery assessing lower extremity function: association with self-reported disability and prediction of mortality and nursing home admission. J Gerontol. 1994;49(2):M85–94. doi:10.1093/geronj/49.2.m85

2. Kempen GI, Steverink N, Ormel J, Deeg DJ. The assessment of ADL among frail elderly in an interview survey: self-report versus performance-based tests and determinants of discrepancies. J Gerontol B Psychol Sci Soc Sci. 1996;51(5):P254–260. doi:10.1093/geronb/51b.5.p254

3. Bean JF, Ölveczky DD, Kiely DK, LaRose SI, Jette AM. Performance-Based Versus Patient-Reported Physical Function: What Are the Underlying Predictors? Phys Ther. 2011;91(12):1804–1811. doi:10.2522/ptj.20100417

4. Alexandre T da S, Corona LP, Nunes DP, Santos JLF, Duarte YA de O, Lebrão ML. Disability in instrumental activities of daily living among older adults: gender differences. Rev Saude Publica. 2014;48(3):379–389. doi:10.1590/s0034-8910.2014048004754

5. Wennie Huang WN, Perera S, VanSwearingen J, Studenski S. Performance measures predict onset of activity of daily living difficulty in community-dwelling older adults. J Am Geriatr Soc. 2010;58(5):844–852. doi:10.1111/j.1532-5415.2010.02820.x

6. den Ouden MEM, Schuurmans MJ, Arts IEMA, van der Schouw YT. Physical performance characteristics related to disability in older persons: a systematic review. Maturitas. 2011;69(3):208–219. doi:10.1016/j.maturitas.2011.04.008

7. Beauchamp MK, Jette AM, Ward RE, et al. Predictive validity and responsiveness of patient-reported and performance-based measures of function in the Boston RISE study. J Gerontol A Biol Sci Med Sci. 2015;70(5):616–622. doi:10.1093/gerona/glu227

8. Mayhew AJ, Griffith LE, Gilsing A, Beauchamp MK, Kuspinar A, Raina P. The Association Between Self-Reported and Performance-Based Physical Function With Activities of Daily Living Disability in the Canadian Longitudinal Study on Aging. J Gerontol A Biol Sci Med Sci. 2020;75(1):147–154. doi:10.1093/gerona/glz122

9. Ferrucci L, Cooper R, Shardell M, Simonsick EM, Schrack JA, Kuh D. Age-Related Change in Mobility: Perspectives From Life Course Epidemiology and Geroscience. J Gerontol A Biol Sci Med Sci. 2016;71(9):1184–1194. doi:10.1093/gerona/glw043

10. Dodds RM, Syddall HE, Cooper R, et al. Grip strength across the life course: normative data from twelve British studies. PloS One. 2014;9(12):e113637. doi:10.1371/journal.pone.0113637

11. Alexander NB. Gait disorders in older adults. J Am Geriatr Soc. 1996;44(4):434–451. doi:10.1111/j.1532-5415.1996.tb06417.x

12. Peeters G, Dobson AJ, Deeg DJH, Brown WJ. A life-course perspective on physical functioning in women. Bull World Health Organ. 2013;91(9):661–670. doi:10.2471/BLT.13.123075

13. Murtagh KN, Hubert HB. Gender differences in physical disability among an elderly cohort. Am J Public Health. 2004;94(8):1406–1411. doi:10.2105/ajph.94.8.1406

14. Kelly-Hayes M, Jette AM, Wolf PA, D’Agostino RB, Odell PM. Functional limitations and disability among elders in the Framingham Study. Am J Public Health. 1992;82(6):841–845. doi:10.2105/ajph.82.6.841

15. Hoeymans N, Feskens EJ, van den Bos GA, Kromhout D. Measuring functional status: cross-sectional and longitudinal associations between performance and self-report (Zutphen Elderly Study 1990-1993). J Clin Epidemiol. 1996;49(10):1103–1110. doi:10.1016/0895-4356(96)00210-7

16. Latham NK, Mehta V, Nguyen AM, et al. Performance-based or self-report measures of physical function: which should be used in clinical trials of hip fracture patients? Arch Phys Med Rehabil. 2008;89(11):2146–2155. doi:10.1016/j.apmr.2008.04.016

17. Katz S, Ford AB, Moskowitz RW, Jackson BA, Jaffe MW. STUDIES OF ILLNESS IN THE AGED. THE INDEX OF ADL: A STANDARDIZED MEASURE OF BIOLOGICAL AND PSYCHOSOCIAL FUNCTION. JAMA. 1963;185:914–919. doi:10.1001/jama.1963.03060120024016

18. Lawton MP, Brody EM. Assessment of older people: self-maintaining and instrumental activities of daily living. The Gerontologist. 1969;9(3):179–186.

19. Ensrud KE, Nevitt MC, Yunis C, et al. Correlates of impaired function in older women. J Am Geriatr Soc. 1994;42(5):481–489. doi:10.1111/j.1532-5415.1994.tb04968.x

20. Studenski S, Perera S, Patel K, et al. Gait speed and survival in older adults. JAMA. 2011;305(1):50–58. doi:10.1001/jama.2010.1923

21. Nelson HD, Nevitt MC, Scott JC, Stone KL, Cummings SR. Smoking, alcohol, and neuromuscular and physical function of older women. Study of Osteoporotic Fractures Research Group. JAMA. 1994;272(23):1825–1831. doi:10.1001/jama.1994.03520230035035

22. Csuka M, McCarty DJ. Simple method for measurement of lower extremity muscle strength. Am J Med. 1985;78(1):77–81. doi:10.1016/0002-9343(85)90465-6

23. Bohannon RW. Test-retest reliability of the five-repetition sit-to-stand test: a systematic review of the literature involving adults. J Strength Cond Res. 2011;25(11):3205–3207. doi:10.1519/JSC.0b013e318234e59f

24. Huntley AL, Johnson R, Purdy S, Valderas JM, Salisbury C. Measures of multimorbidity and morbidity burden for use in primary care and community settings: a systematic review and guide. Ann Fam Med. 2012;10(2):134–141. doi:10.1370/afm.1363

25. Andersen P, Gill R. Cox’s Regression Model for Counting Processes: A Large Sample Study. Ann Stat. 1982;10(4):110–1120. doi:10.1214/aos/1176345976

26. Lee MC, Hsu CC, Tsai YF, Chen CY, Lin CC, Wang CY. Criterion-Referenced Values of Grip Strength and Usual Gait Speed Using Instrumental Activities of Daily Living Disability as the Criterion. J Geriatr Phys Ther 2001. 2018;41(1):14–19. doi:10.1519/JPT.0000000000000106

27. Guralnik JM, Ferrucci L, Pieper CF, et al. Lower extremity function and subsequent disability: consistency across studies, predictive models, and value of gait speed alone compared with the short physical performance battery. J Gerontol A Biol Sci Med Sci. 2000;55(4):M221–231. doi:10.1093/gerona/55.4.m221

28. Onder G, Penninx BWJH, Ferrucci L, Fried LP, Guralnik JM, Pahor M. Measures of physical performance and risk for progressive and catastrophic disability: results from the Women’s Health and Aging Study. J Gerontol A Biol Sci Med Sci. 2005;60(1):74–79. doi:10.1093/gerona/60.1.74

29. Seino S, Yabushita N, Kim M ji, et al. Comparison of a combination of upper extremity performance measures and usual gait speed alone for discriminating upper extremity functional limitation and disability in older women. Arch Gerontol Geriatr. 2012;55(2):486–491. doi:10.1016/j.archger.2011.10.011

30. Cesari M, Onder G, Zamboni V, et al. Physical function and self-rated health status as predictors of mortality: results from longitudinal analysis in the ilSIRENTE study. BMC Geriatr. 2008;8:34. doi:10.1186/1471-2318-8-34

31. Rolland Y, Lauwers-Cances V, Cesari M, Vellas B, Pahor M, Grandjean H. Physical performance measures as predictors of mortality in a cohort of community-dwelling older French women. Eur J Epidemiol. 2006;21(2):113–122. doi:10.1007/s10654-005-5458-x

32. Blain H, Carriere I, Sourial N, et al. Balance and walking speed predict subsequent 8-year mortality independently of current and intermediate events in well-functioning women aged 75 years and older. J Nutr Health Aging. 2010;14(7):595–600. doi:10.1007/s12603-010-0111-0

33. Gérin-Lajoie M, Richards CL, McFadyen BJ. The circumvention of obstacles during walking in different environmental contexts: a comparison between older and younger adults. Gait Posture. 2006;24(3):364–369. doi:10.1016/j.gaitpost.2005.11.001

34. Buchner DM, Larson EB, Wagner EH, Koepsell TD, de Lateur BJ. Evidence for a non-linear relationship between leg strength and gait speed. Age Ageing. 1996;25(5):386–391. doi:10.1093/ageing/25.5.386

35. Ostchega Y, Dillon CF, Lindle R, Carroll M, Hurley BF. Isokinetic leg muscle strength in older americans and its relationship to a standardized walk test: data from the national health and nutrition examination survey 1999-2000. J Am Geriatr Soc. 2004;52(6):977–982. doi:10.1111/j.1532-5415.2004.52268.x

36. Buchman AS, Shah RC, Leurgans SE, Boyle PA, Wilson RS, Bennett DA. Musculoskeletal pain and incident disability in community-dwelling older adults. Arthritis Care Res. 2010;62(9):1287–1293. doi:10.1002/acr.20200

37. Persad CC, Jones JL, Ashton-Miller JA, Alexander NB, Giordani B. Executive function and gait in older adults with cognitive impairment. J Gerontol A Biol Sci Med Sci. 2008;63(12):1350–1355. doi:10.1093/gerona/63.12.1350

38. Watson NL, Rosano C, Boudreau RM, et al. Executive function, memory, and gait speed decline in well-functioning older adults. J Gerontol A Biol Sci Med Sci. 2010;65(10):1093–1100. doi:10.1093/gerona/glq111

39. Pérez-Zepeda MU, González-Chavero JG, Salinas-Martinez R, Gutiérrez-Robledo LM. RISK FACTORS FOR SLOW GAIT SPEED: A NESTED CASE-CONTROL SECONDARY ANALYSIS OF THE MEXICAN HEALTH AND AGING STUDY. J Frailty Aging. 2015;4(3):139–143. doi:10.14283/jfa.2015.63

40. Fritz S, Lusardi M. White paper: “walking speed: the sixth vital sign.” J Geriatr Phys Ther 2001. 2009;32(2):46–49.

41. Middleton A, Fritz SL, Lusardi M. Walking speed: the functional vital sign. J Aging Phys Act. 2015;23(2):314–322. doi:10.1123/japa.2013-0236

42. Wolinsky FD, Bentler SE, Hockenberry J, et al. Long-term declines in ADLs, IADLs, and mobility among older Medicare beneficiaries. BMC Geriatr. 2011;11:43. doi:10.1186/1471-2318-11-43

43. Lyons JG, Heeren T, Stuver SO, Fredman L. Assessing the agreement between 3-meter and 6-meter walk tests in 136 community-dwelling older adults. J Aging Health. 2015;27(4):594–605. doi:10.1177/0898264314556987

44. Sallinen J, Stenholm S, Rantanen T, Heliövaara M, Sainio P, Koskinen S. Hand-grip strength cut points to screen older persons at risk for mobility limitation. J Am Geriatr Soc. 2010;58(9):1721–1726. doi:10.1111/j.1532-5415.2010.03035.x

45. Bohannon RW. Reference values for the five-repetition sit-to-stand test: a descriptive meta-analysis of data from elders. Percept Mot Skills. 2006;103(1):215–222. doi:10.2466/pms.103.1.215-222

